# COVID-19 Bed Surge and Rationing of Rehabilitation Services in Japan: A Seasonal Autoregressive Integrated Moving Average (SARIMA) Analysis of Care Utilization with 10 years claims data

**DOI:** 10.1101/2025.06.18.25329839

**Authors:** Yuki Egashira, Ryo Watanabe

## Abstract

**Objective:** To explore the effect of COVID-19 and associated medical resource prioritization called “rationing” on the provision of rehabilitation services in Japan, focusing on cerebrovascular, musculoskeletal, and respiratory rehabilitation in the pre-COVID-19, during-COVID-19, and post-COVID-19 periods.

**Design:** Retrospective study using seasonal autoregressive integrated moving average model to predict expected values, which were compared with actual values to calculate observed-to-expected ratios.

**Setting:** Acute care hospitals in Kanagawa Prefecture, which has the second largest population in Japan after Tokyo, covering April 2014 to March 2024.

**Participants:** Patients aged 0–74 years who were enrolled in the National Health Insurance of Kanagawa Prefecture and underwent the studied types of rehabilitation.

**Exposure:** COVID-19 pandemic waves and associated bed utilization rates, with multiple distinct peaks.

**Outcome measures:** The difference between the predicted and actual values of the volume of rehabilitation services provided and the number of patients per insured person calculated as observed-to-expected ratios in response to the peak of bed utilization against COVID-19.

**Results:** The observed-to-expected ratio of inpatient rehabilitation services for cerebrovascular showed a significant decrease after five waves at −14.3%, and musculoskeletal conditions showed a similar decline during periods of high COVID-19 bed utilization. Outpatient services experienced sharp declines initially but showed differential recovery patterns. Respiratory rehabilitation displayed unique patterns, with inpatient services increasing up to 62.4% above expected levels until September 2021, before sharply declining. By March 2024, musculoskeletal rehabilitation demonstrated complete recovery, cerebrovascular rehabilitation showed partial recovery, while respiratory rehabilitation exhibited mixed patterns with persistent outpatient deficits.

**Conclusions:** The COVID-19 pandemic significantly impacted rehabilitation services in Japan, with inpatient services for cerebrovascular and musculoskeletal conditions being particularly vulnerable to disruptions during high COVID-19 bed utilization periods. The differential recovery patterns across rehabilitation types, with some structural changes persisting beyond the acute pandemic phase, indicate the need for flexible healthcare systems to deal with future healthcare crises. These findings underscore the importance of developing strategies to maintain essential rehabilitation services during public health emergencies, especially considering the aging global population and rising demand for rehabilitation.

**Article Summary:** *Strengths and Limitations of this study:*

- This study is the first to use 10 years of large-scale claims data (Apr 2014–Mar 2024) to show how COVID-19 bed surges disrupted rehabilitation services across multiple waves, including the post-Category 5 period.
- The study findings indicate that service disruption varied by type: cerebrovascular and musculoskeletal rehab were more vulnerable, while respiratory rehab showed unique patterns of increase during the pandemic.
- A Seasonal Autoregressive Integrated Moving Average model using 72 months of pre-COVID data enabled accurate prediction of expected service levels across pre-, during-, and post-pandemic periods.
- The study design could not identify the causal relationship between healthcare resource constraints and changes in patient care-seeking behaviors as primary drivers of decreased service utilization.
- The study focused on a single prefecture, which may limit generalizability to other regions with different healthcare infrastructure.

## Background

The coronavirus disease 2019 (COVID-19) pandemic, declared by the World Health Organization (WHO) on March 11, 2020, has substantially affected various health services globall.^1^ Although the WHO declared the end of the pandemic in May 2023, especially at the initial phase of the pandemic, many countries implemented lockdowns to mitigate the spread of the pandemic. With limited medical resources, non-urgent surgeries were canceled or delayed, patient numbers were restricted, and rehabilitation beds were repurposed for COVID-19, except for urgent rehabilitation cases.^2^ These measures, known as “rationing,” involve allocating limited healthcare resources to those who can potentially benefit from treatment.^3^

Several studies have shown a decrease in elective surgeries during the COVID-19 pandemic.^4–9^ However, studies of the impact of COVID-19 on rehabilitation within healthcare delivery systems are limited. Rehabilitation is crucial, as it will increase the number of patients worldwide, prove cost-effective, and improve the patient’s function and quality of life. A global study found that the demand for rehabilitation increased by 63% in 2019 compared to 1990, driven primarily by population aging, particularly among those aged 50–70 years.^10^ The World Health Organization estimates that the global population over 60 years will increase from 12% to 22% between 2015 and 2050.^11^

In Japan, which has one of the highest aging rates globally, national plans include a strategy for converting acute care beds into post-acute care beds to secure hospital beds with rehabilitation functions.^12^ The rapid increase in the demand for post-acute beds in Japan implies that other countries with aging populations may face similar situations of requiring post-acute beds and medical care. Additionally, the economic benefits of rehabilitation are evident. Systematic reviews of economic evaluations show that rehabilitation is cost-effective, achieving high clinical outcomes at a low cost.^13^

Rehabilitation contributes to the functional improvement of targeted areas and maintenance and improvement of the quality of life. For example, it improves muscle strength, reduces pain, and enhances the quality of life in patients with musculoskeletal dysfunction.^14^ ^15^ Similarly, respiratory rehabilitation improves respiratory function and quality of life in patients with interstitial lung and chronic obstructive pulmonary diseases.^16–18^ In patients with stroke, early rehabilitation can improve functional transitions and shorten hospitalization.^19^ ^20^ However, various factors have influenced the implementation of different types of rehabilitation during the COVID-19 pandemic. For cerebrovascular disease rehabilitation, changes in treatment protocols, closure of rehabilitation units, and lack of staff and protective equipment have affected the healthcare delivery system.^21–23^ Conversely, the demand for respiratory rehabilitation has increased because it can improve the prognosis of COVID-19 sequelae.^24^

Previous studies had several limitations that must be overcome. First, the status of medical provisions for rehabilitation varies by type, and cross-site studies are limited. Second, most studies focused on the early stages of the pandemic and did not examine long-term trend changes including post-COVID-19. Therefore, we used long-term claims data, including data from the initial phase of COVID-19. In this study, we hypothesized that the volume of rehabilitation services provided and the decrease in patient numbers resulted from increased bed utilization caused by the upsurge of the bed against COVID-19. Additionally, we assumed that an increase in constraints of the beds due to COVID-19 may have led to certain difficulties in the provision of rehabilitation care, implying the occurrence of rationing.^25^ This study aimed to examine the impact of COVID-19 and the associated “rationing” of medical resources on the provision of rehabilitation services in Japan in the pre-COVID-19, during-COVID-19, and post-COVID-19 periods.

## Methods

### Study Design and Setting

We conducted a retrospective study using claims data from Kanagawa Prefecture, Japan, covering the time frame from April 2014 to March 2024. Our study was divided in three periods: pre-COVID-19, during COVID-19, and post-COVID-19.

We selected Japan for this study because it is one of the first countries to have an aging population, and a significant impact of COVID-19 on rehabilitation services was anticipated.^26^ However, studies on quantitative and long-term effects are limited.^27^ Kanagawa Prefecture was selected for three primary reasons. First, it has the second largest population in Japan after Tokyo, ensuring high representativeness. Second, it has the lowest number of hospital beds per capita in Japan,^28^ making it particularly sensitive to bed shortages. Third, previous studies have not fully addressed the impact of the December 2020–January 2021 outbreak and the effects of Delta and Omicron strains on medical resources.^27^ This study focused on three types of rehabilitation: cerebrovascular, musculoskeletal, and respiratory.

### Data Source and Participants

We utilized the Kokuho Database System (KDB), one of the largest databases in Japan, which contains claims data for Kanagawa Prefecture. The KDB has been used in previous clinical and epidemiological studies.^29^ ^30^ The claims data included age, sex, place of residence, medical service codes, disease codes, quantity and frequency of services, and unique de-identified patient codes.

The participants were patients aged 0–74 years who were enrolled in the National Health Insurance of Kanagawa Prefecture between April 2014 and March 2024 and received rehabilitation (cerebrovascular, musculoskeletal, or respiratory). The National Health Insurance is a public insurance program for private employers, nonworkers, and retirees, covering approximately 18.0% of the population aged 0–64 years and 68.5% of the population aged 65–74 years.^31^ To examine the impact of hospital bed shortage, we specifically included patients who received treatment at acute care hospitals in Kanagawa Prefecture. We defined acute hospitals as those that claimed a fee for acute hospitals during the study period.

### Outcome Measures

The primary outcome was the difference between the predicted and actual values of the volume of rehabilitation services provided and the number of patients per insured person after April 2020 following the COVID-19 outbreak. These values were calculated separately for inpatient and outpatient services. In Japan, one rehabilitation unit is claimed for each defined hour of rehabilitation. We defined the monthly volume as the total number of units claimed for each month.

We descriptively observed the relationship between the peak periods of bed utilization and outcomes during several waves of the pandemic: (1st: 2020/4 first state of emergency in Japan, 3rd: 2021/1 third wave, first bed shortage, 5th: 2021/8 Delta strain outbreak, 6th: 2022/2 Omicron outbreak, 7th: 2022/8, and 8th: 2023/1). The data were obtained from the Ministry of Labor and Welfare, and the peak utilization rate for each pandemic wave was analyzed.^32^

### Statistical Analysis

We employed a seasonal autoregressive integrated moving average (SARIMA) model for our analysis. The SARIMA comprises an autoregressive component that predicts the future using historical data, a difference component that removes data trends, a moving average component that predicts the future using historical forecast errors, and a seasonal component that accounts for seasonal patterns.

The SARIMA model is represented as (p, d, q) × (P, D, Q, m), where p, d, and q are the parameters of the ARIMA model, and P, D, Q, and m are the parameters of the seasonal component, with m being the period of seasonality. We estimated trends for 72 months, without missing data, from April 2014 to March 2020 regarding the volume of rehabilitation units provided and the number of patients per million insured persons in each rehabilitation program. Because the number of insured individuals were publicly available only through FY2022, we used the FY2014–FY2022 average percentage change for FY2023 values (April 2023–March 2024).^33^ The model selection procedure involved four steps. First, we used the Dickey–Fuller test as a unit root test to assess stationarity, with a significance level of p <0.05. Second, the parameters of the SARIMA model were set using Stata’s Arimaauto package, which selects the model with the lowest Akaike Information Criterion among the candidate parameters.^34^ Third, the selected models were subjected to a white noise test using the Ljung–Box chi-square test, with a significance level of p >0.05. If the white noise test result was below this value, the parameters were reset, and the procedure was repeated until the result exceeded 0.05. Finally, to evaluate the fit of the final selected model, we calculated the mean absolute percentage error using the values of the forecast model and actual measurements from April 2016 to March 2020. Municipalities with missing data were excluded.

For most ARIMA models, confidence intervals were calculated using Stata’s stdp option, which provides theoretical standard errors accounting for parameter uncertainty. For complex models where stdp was unavailable (particularly with moving average components), we used residual-based standard deviation (square root of sum of squared residuals divided by degrees of freedom), similar to approaches employed in healthcare utilization forecasting studies when theoretical standard errors are unavailable. The residual-based method produces wider, more conservative confidence intervals than stdp as it does not account for precision gains from parameter estimation. Both methods constructed 95% confidence intervals as predicted value ± 1.96 × standard error, with lower bounds constrained to zero. All statistical analyses were conducted using STATA, MP16.1.

### Patient and public involvement

Patients and/or the public were not involved in the design, conduct, reporting, or dissemination plans of this research.

## Results

After excluding eight municipalities with missing data, the analysis included data from 48 municipalities to ensure a comprehensive and consistent dataset throughout the study period. Figure 1 illustrates trends in the utilization of COVID-19 beds in Kanagawa Prefecture. The graph reveals multiple distinct peaks across the study period: January 2021 (third wave) at 47% utilization, August 2021 (fifth wave) at 72% utilization, February 2022 (sixth wave) at 65% utilization, September 2022 (seventh wave) at 88% utilization, and January 2023 (eighth wave) at 78% utilization. These peaks correspond to major surges in COVID-19 cases and provide context for the changes in the provision of rehabilitation services. The seventh wave in September 2022 showed the highest bed utilization rate at 88%, indicating the most severe strain on the healthcare system throughout the entire observation period.

**Figure 1.**
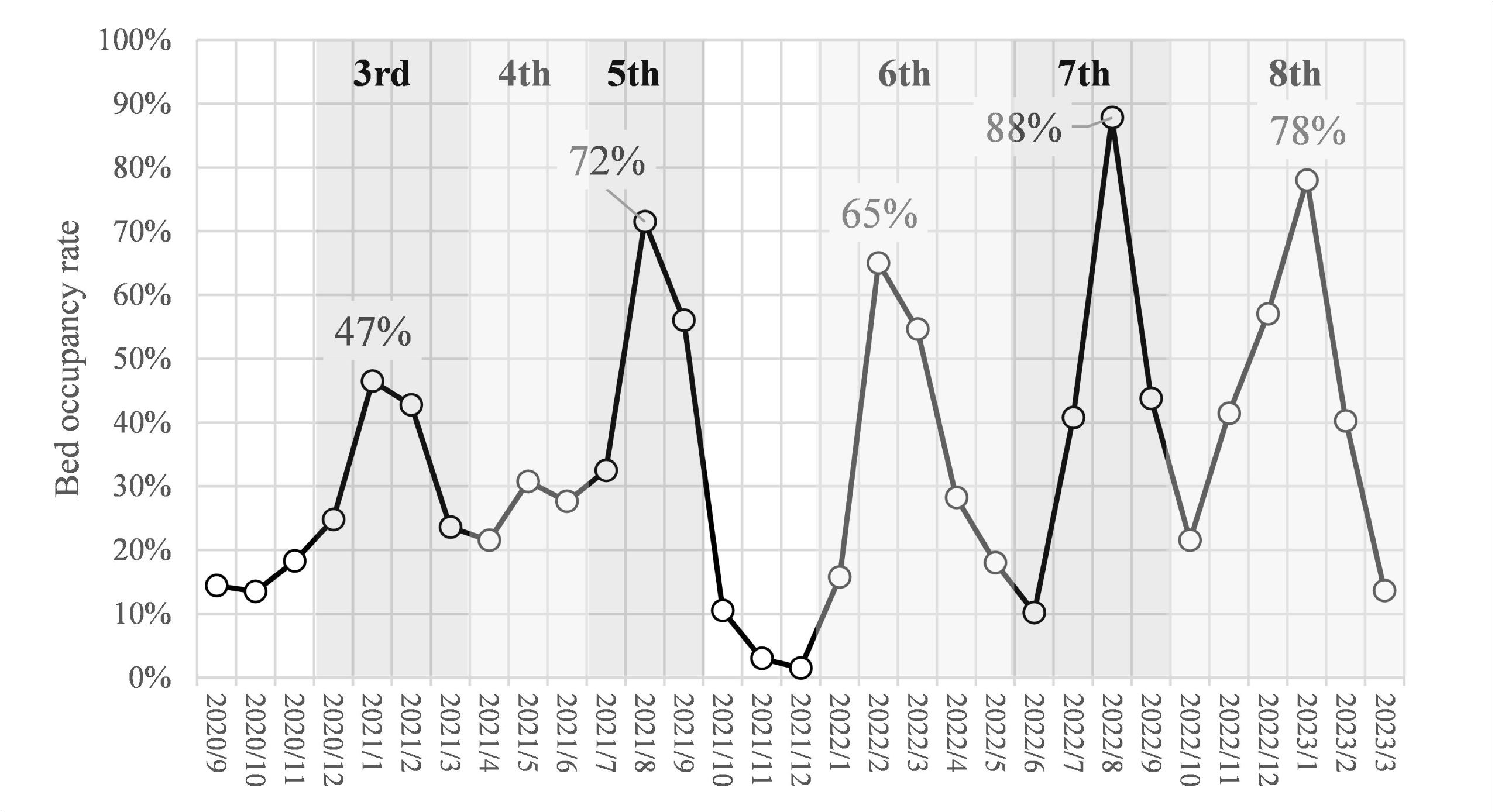
COVID-19 bed utilization trends.

### Distribution of Rehabilitation Services

Table 1 presents the gender distribution across different rehabilitation services during the three study periods. For cerebrovascular rehabilitation, males consistently comprised approximately 61% of inpatient cases and 57-58% of outpatient cases across all periods, with no statistically significant changes observed (p=0.12 for inpatient, p=0.08 for outpatient). Musculoskeletal rehabilitation showed a contrasting pattern, with females representing the majority in both settings: approximately 59% of inpatient cases and 65% of outpatient cases, remaining stable throughout the study periods (p=0.34 for inpatient, p=0.15 for outpatient). Respiratory rehabilitation demonstrated a significant shift in gender distribution for inpatient services, where the proportion of males decreased from 67.8% before COVID-19 to 65.9% in the post-COVID-19 period (p=0.02), while outpatient services showed fluctuating patterns without statistical significance (p=0.18).

**Table 1.**
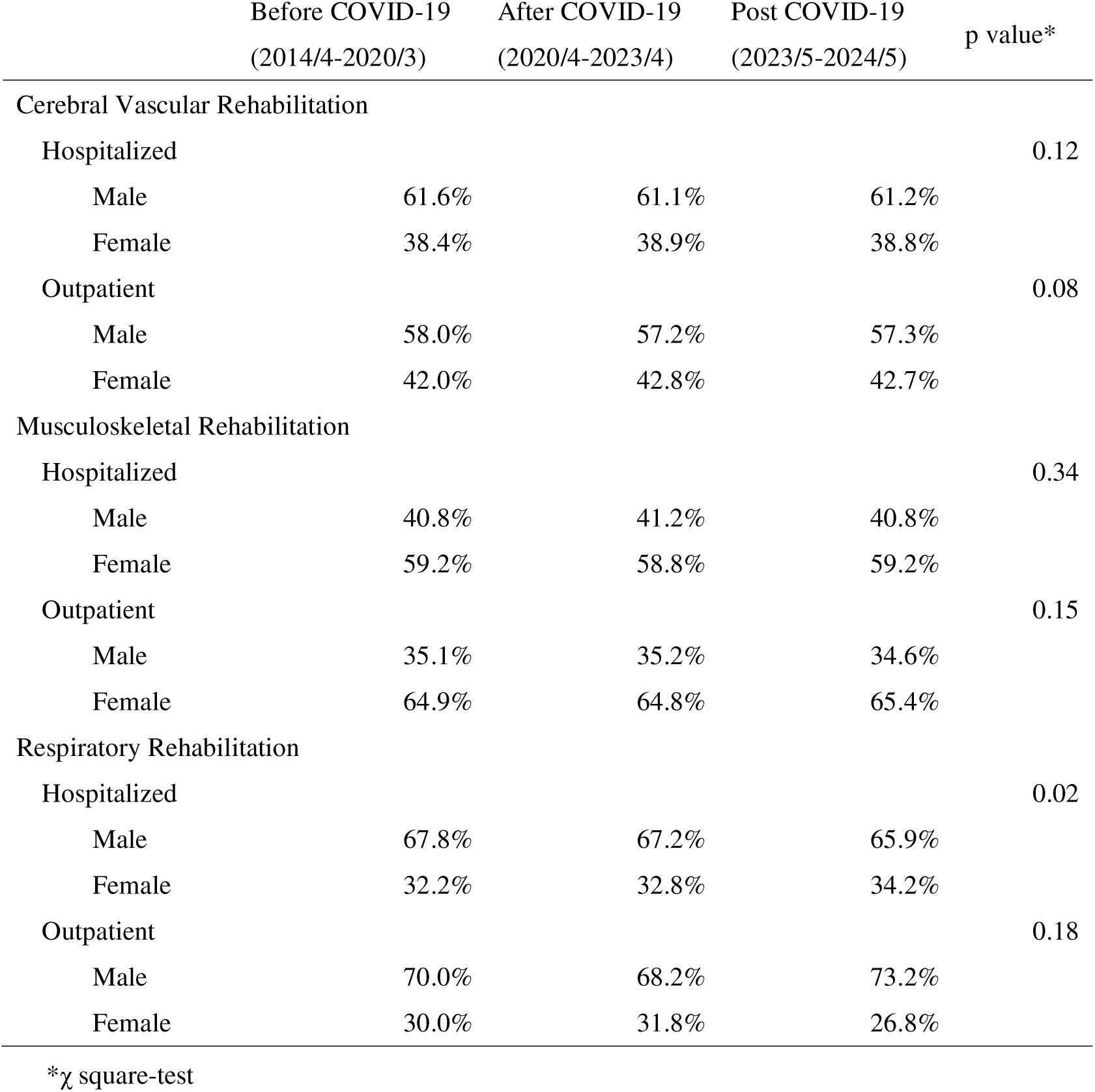
Gender distribution by rehabilitation services.

Table 2 illustrates the age distribution changes across the study periods. Cerebrovascular rehabilitation patients showed a notable trend toward younger ages, with inpatient mean ages decreasing from 63.4 years before COVID-19 to 62.1 years post-COVID-19, and outpatient mean ages declining more substantially from 48.9 to 44.0 years (both p<0.01). Conversely, musculoskeletal rehabilitation patients demonstrated a gradual aging trend, with inpatient mean ages increasing from 64.5 to 65.5 years and outpatient mean ages rising from 61.7 to 62.9 years (both p<0.01). Respiratory rehabilitation patients maintained the highest mean ages across all categories (66-68 years), with minimal but statistically significant changes over time (both p<0.01).

**Table 2.** Age distribution by rehabilitation services.

|  | Mean (SD <sup>*1</sup>) |  |  |  |  |  | p value <sup>*2</sup> |
| --- | --- | --- | --- | --- | --- | --- | --- |
|  | Before COVID-19 |  | After COVID-19 |  | Post COVID-19 |  |  |
|  | (2014/4-2020/3) |  | (2020/4-2023/4) |  | (2023/5-2024/5) |  |  |
| Cerebral Vascular Rehabilitation |  |  |  |  |  |  |  |
| Hospitalized | 63.4 | (12.3) | 63.0 | (12.6) | 62.1 | (13.0) | <0.01 |
| Outpatient | 48.9 | (23.7) | 45.6 | (24.5) | 44.0 | (24.4) | <0.01 |
| Musculoskeletal Rehabilitation |  |  |  |  |  |  |  |
| Hospitalized | 64.5 | (11.9) | 65.4 | (11.3) | 65.5 | (11.0) | <0.01 |
| Outpatient | 61.7 | (14.4) | 62.4 | (13.9) | 62.9 | (13.3) | <0.01 |
| Respiratory Rehabilitation |  |  |  |  |  |  |  |
| Hospitalized | 66.2 | (11.4) | 66.4 | (10.7) | 66.9 | (11.0) | <0.01 |
| Outpatient | 67.1 | (11.1) | 68.2 | (8.3) | 67.7 | (9.2) | <0.01 |

\*1 SD: Standard Deviation
\*2 ANOVA-test

Table 3 presented the number of units and percentage of patients by rehabilitation type during the study period. For cerebrovascular rehabilitation, 95.4% of the units provided inpatient care and 4.6% provided outpatient care; 68.4% of the patients received inpatient care, and 31.6% received outpatient care. Musculoskeletal rehabilitation showed a different pattern, where 69.5% of the units provided inpatient services and 30.5% provided outpatient services; however, 31.1% of the patients received inpatient care and 68.9% received outpatient care. Respiratory rehabilitation was predominantly inpatient, with 96.3% of the units and 88.1% of the patients in inpatient settings compared to 3.7% of the units and 11.9% of the patients in outpatient settings.

**Table 3.**
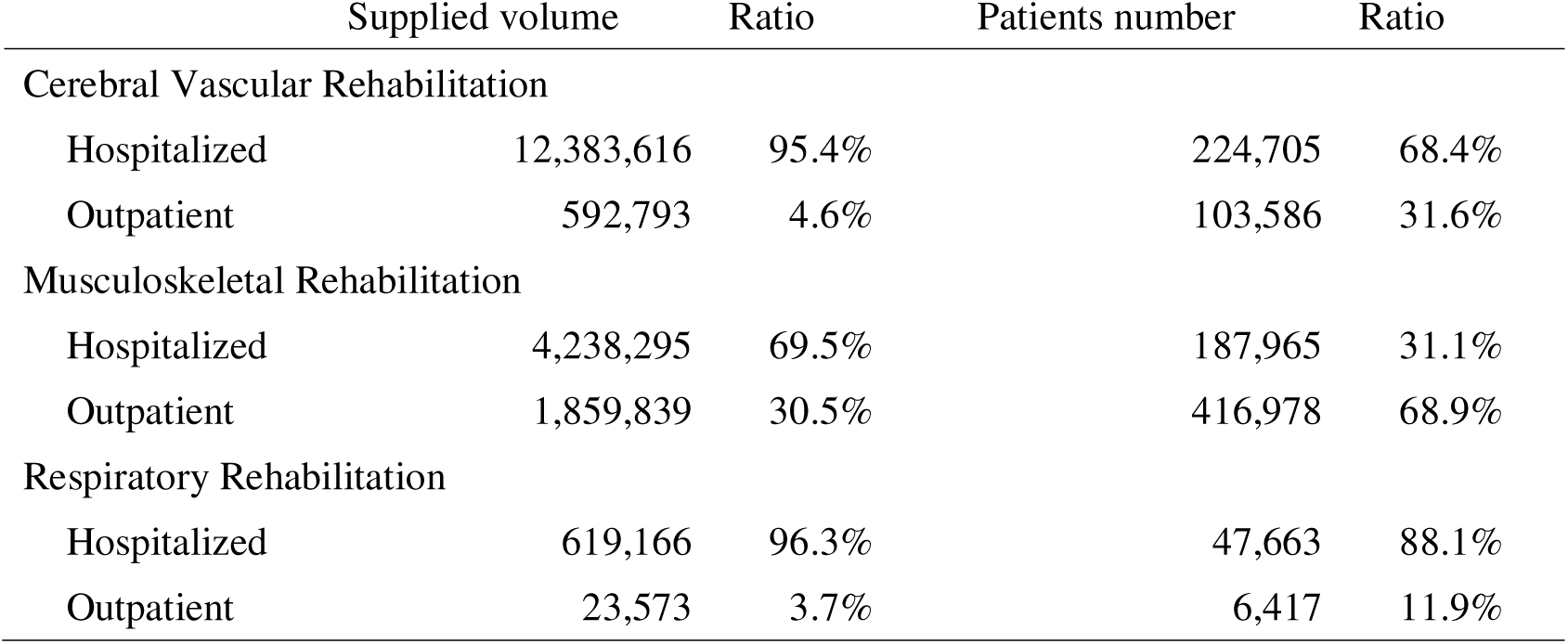
Supplied volume and number of patients.

These figures highlighted the predominance of inpatient services for cerebrovascular and respiratory rehabilitation in terms of service units and patient numbers. Conversely, musculoskeletal rehabilitation showed a more balanced distribution, with a higher proportion of outpatient patients despite more inpatient service units.

Table 4 depicts the average number of monthly units per claim across the three study periods. For cerebrovascular rehabilitation, inpatient services showed an initial increase from 54.9 units before COVID-19 to 56.2 units during the COVID-19 period, followed by a decrease to 53.1 units post-COVID-19. Outpatient services demonstrated a consistent declining trend from 5.8 to 5.1 units across the periods. Musculoskeletal rehabilitation exhibited substantial increases in inpatient services, rising from 21.0 units before COVID-19 to 25.4 units during COVID-19, with a slight decrease to 24.6 units post-COVID-19 (overall increase of 17.1%). Outpatient services showed modest but consistent growth from 4.4 to 4.7 units (6.8% increase). Respiratory rehabilitation demonstrated the most pronounced changes, with inpatient services increasing from 12.1 units before COVID-19 to 14.3 units during COVID-19, then declining to 13.5 units post-COVID-19 (overall increase of 11.6%). Outpatient services showed initial decline from 3.8 to 3.2 units during COVID-19, followed by recovery to 4.1 units post-COVID-19.

**Table 4.**
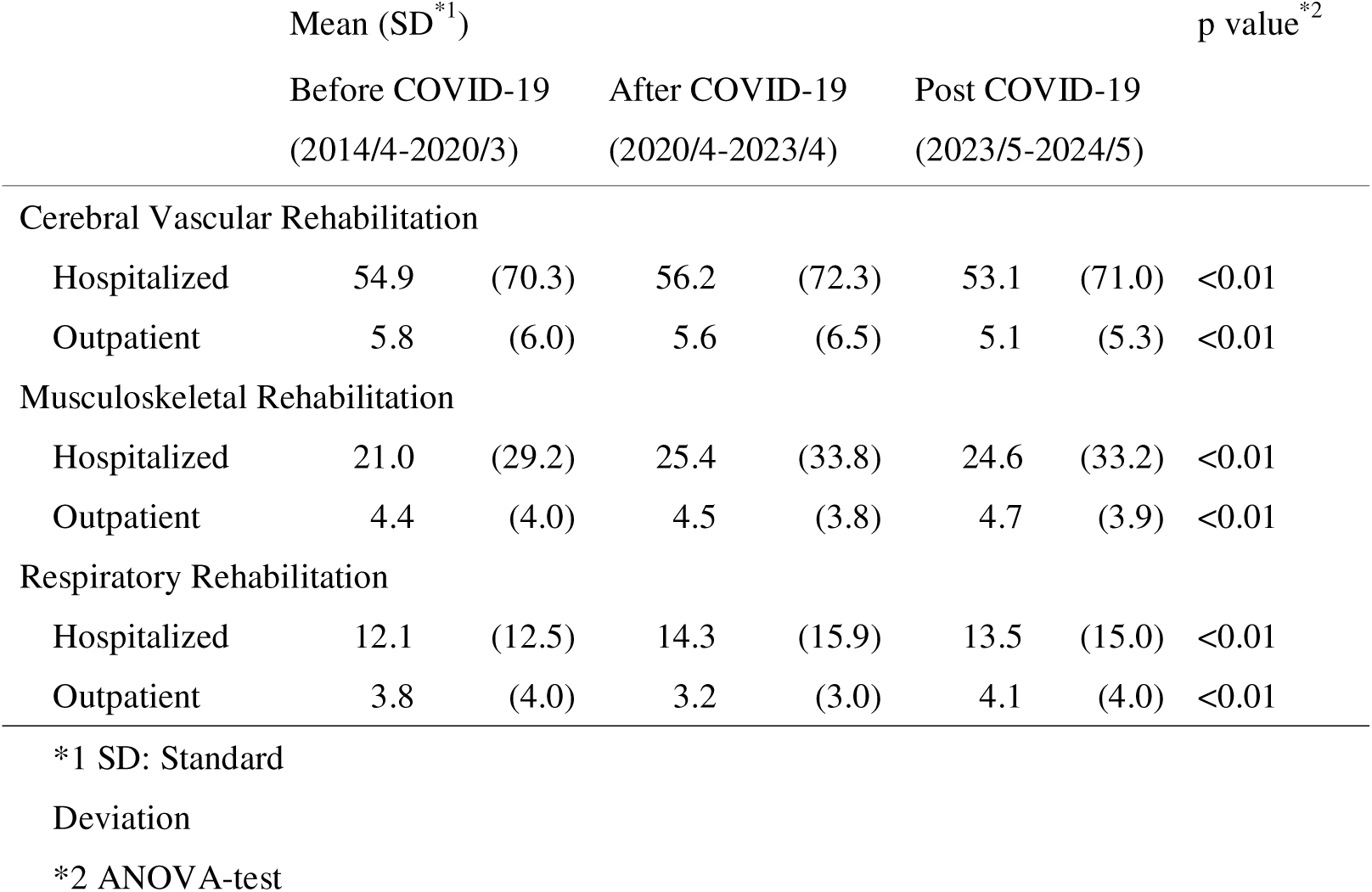
Changes in Rehabilitation Service Provision.

These results indicate that the COVID-19 pandemic had lasting impacts on rehabilitation service delivery patterns, with inpatient services generally showing increased intensity while outpatient services demonstrated more variable responses across different rehabilitation types.

### Observed-to-Expected Ratios

#### Observed-to-Expected Ratios During COVID-19 and Recovery Period

Figures 2 and 3 present the temporal evolution of rehabilitation service utilization through observed-to-expected (OE) ratios, revealing distinct patterns across rehabilitation types and settings. The analysis covered the complete period, encompassing the pre-pandemic baseline, the entire COVID-19 pandemic trajectory, the transition to Category 5 classification in May 2023, and the subsequent recovery phase.

**Figure 2.**
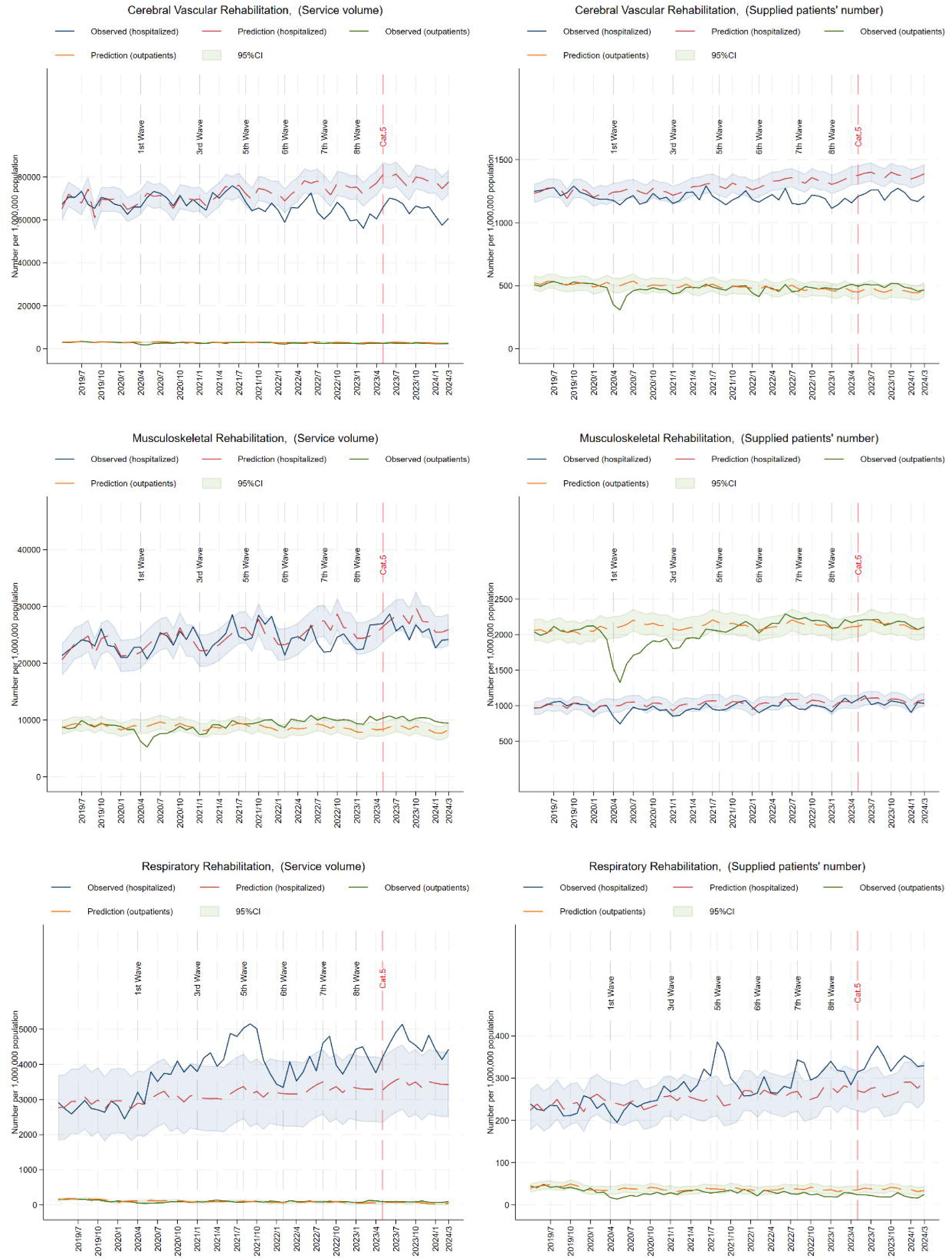
Observed and predicted number of supplied services and patients.

**Figure 3.**
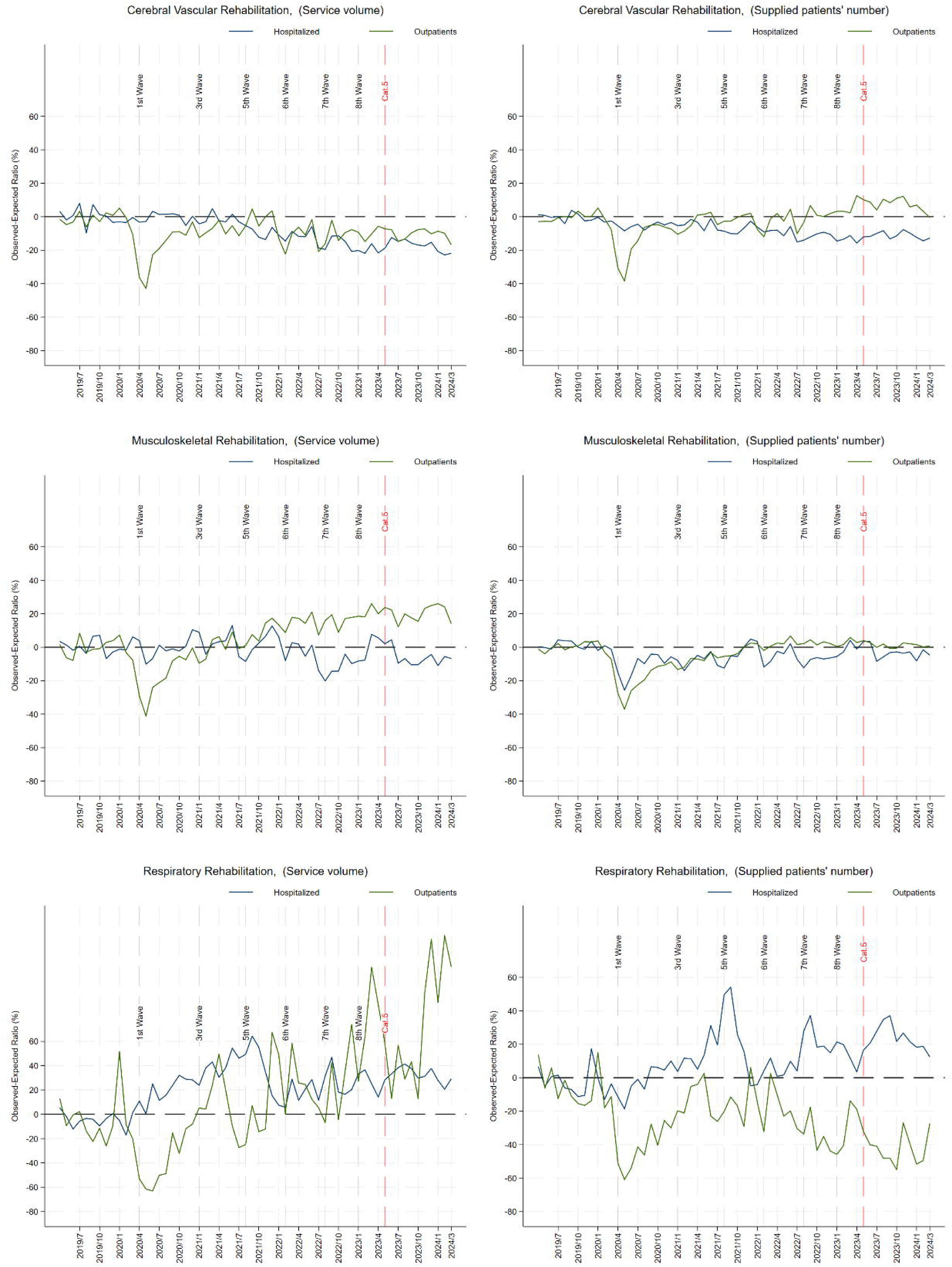
Observed-to-expected ratio.

#### Cerebrovascular Rehabilitation

Cerebrovascular rehabilitation services demonstrated moderate initial resilience but sustained negative impacts throughout most of the observation period. Inpatient unit provision exhibited relative stability with OE ratios fluctuating around ±5.0% until August 2021 (fifth wave), before experiencing a notable decline to −13.6% between the end of the fifth wave and the start of the sixth wave in early 2022. Following the Category 5 reclassification in May 2023, inpatient units showed gradual stabilization but remained below expected levels through March 2024. Inpatient patient numbers demonstrated more pronounced sensitivity to pandemic disruptions, reaching the lowest OE ratio of −14.3%, immediately following the fifth wave and maintaining consistently negative ratios after the initial pandemic onset, with limited recovery evident by March 2024.

Outpatient cerebrovascular services experienced the most severe immediate impact, with OE ratios plummeting to −42.9% for units and −38.5% for patients during the first wave in May 2020. While unit provision gradually improved and temporarily shifted to positive territory in late 2021, it returned to negative values after January 2022 and showed only marginal improvement following the Category 5 transition. Patient numbers demonstrated a more sustained recovery trajectory, turning slightly positive after April 2021, maintaining improvement through 2022–2023, and showing stabilization to neutral levels by early 2024.

#### Musculoskeletal Rehabilitation

Musculoskeletal rehabilitation services showed cyclical patterns that were closely aligned with infection waves, with evidence of systematic recovery by the end of the observation period. Inpatient services recorded notable decreases during peak bed utilization periods, with OE ratios of −9.8% during the first wave (May 2020), −7.9% during the fifth wave (August 2021), and −6.9% during the sixth wave (February 2022). Following the Category 5 reclassification, inpatient units demonstrated clear recovery trends, approaching neutral levels by March 2024. Patient numbers, which reached the lowest point of −25.7% during the first wave, showed robust recovery throughout 2023 and achieved positive OE ratios by early 2024.

Outpatient musculoskeletal services experienced the most dramatic initial disruption, with OE ratios declining to −40.9% for units and −36.3% for patients in May 2020. Recovery patterns showed unit provision transitioning to positive trends by March 2021, maintaining steady improvement through 2024, and achieving OE ratios of approximately +5% to +10% by March 2024. Patient numbers demonstrated similar recovery, achieving sustained positive ratios from September 2021 onwards and stabilizing at positive levels throughout the post-Category 5 period.

#### Respiratory Rehabilitation

Respiratory rehabilitation displayed the most distinctive utilization patterns throughout the observation period. Inpatient services experienced substantial increases beginning in April 2020, likely reflecting COVID-19-related demand for respiratory support services. This surge peaked dramatically in September 2021, reaching OE ratios of +62.4% for units and +53.0% for patients—the highest positive deviations observed across all rehabilitation categories. Following a sharp decline in December 2021, inpatient respiratory services showed volatile patterns through 2022 before demonstrating normalization trends post-Category 5 transition, approaching baseline levels by March 2024.

Outpatient respiratory rehabilitation services demonstrated the most persistent negative trends throughout the observation period. Following severe reductions during the first wave (−71.4% for units in June 2020 and −67.2% for patients in May 2020), these services showed limited recovery through 2022 and 2023. Even after the Category 5 reclassification, outpatient respiratory services remained substantially below expected levels, with OE ratios of approximately −30% to −40% persisting through March 2024, suggesting structural changes in respiratory care delivery patterns.

#### Recovery Trajectories and Post-Category 5 Trends

The transition to Category 5 classification in May 2023 marked a turning point for several rehabilitation services. Musculoskeletal rehabilitation demonstrated the most complete recovery, with both inpatient and outpatient services achieving or exceeding expected levels by March 2024. Cerebrovascular rehabilitation showed partial recovery, with outpatient services approaching baseline levels while inpatient services remained moderately below expectations. Respiratory rehabilitation presented mixed patterns, with inpatient services normalizing but outpatient services showing persistent deficits, suggesting lasting changes in care delivery models.

#### Cross-Category Analysis and Healthcare System Adaptation

The five-year analysis revealed complex, type-specific responses in rehabilitation service provision during the COVID-19 pandemic and recovery period. The data demonstrate that while acute disruptions occurred across all rehabilitation types during 2020–2021, recovery patterns varied substantially. By March 2024, approximately four years after the initial pandemic impact, musculoskeletal rehabilitation had achieved full recovery, cerebrovascular rehabilitation showed partial recovery, and respiratory rehabilitation exhibited mixed but improving trends. These differential recovery patterns suggest that pandemic impacts on rehabilitation services were not uniform and that some structural changes in care delivery may persist beyond the acute pandemic phase.

## Discussion

Our results suggest a strong relationship between COVID-19 bed utilization and decreased cerebrovascular and musculoskeletal rehabilitation services. The lowest OE ratio for inpatient rehabilitation coincided with peak COVID-19 bed utilization after the third wave, except for cerebrovascular rehabilitation during the fifth wave, implying that hospital bed shortages affected the rationing of rehabilitation services. In case of cerebrovascular rehabilitation during the fifth wave, for peak COVID-19 bed utilization, OE ratio also showed a decreasing trend but the lowest timing was observed after the peak COVID-19 bed utilization. Previous studies reported similar decreases in rehabilitation services during the early stages of the pandemic.^35^ ^36^ Our study extends these findings to later periods of high global infection rates and acute hospital bed shortages. The decrease in services may be attributed to infection control measures within hospitals, as a German study found that 15.7% of the surveyed hospitals experienced partial or complete ward closures amid COVID-19 outbreaks.^37^

Inpatient rehabilitation for cerebrovascular and musculoskeletal conditions showed high sensitivity to bed shortages and infection outbreaks. Conversely, outpatient rehabilitation experienced a sharp decline during the first wave but subsequently stabilized. This differential impact is notable, as inpatient rehabilitation accounted for 95.8% and 68.6% of the total volume of cerebrovascular and musculoskeletal services, respectively. These rehabilitation fields often require intensive inpatient care, followed by outpatient rehabilitation. Although rehabilitation may not directly prevent mortality, it substantially affects the patient’s quality of life and subsequent physical function. Ensuring stable provision of inpatient rehabilitation during health crises remains a critical challenge for healthcare systems. Conversely, respiratory rehabilitation showed a consistent upward trend until September 2021, likely reflecting the growing demand because of COVID-19. This trend aligns with those reported in previous studies, demonstrating the effectiveness of respiratory rehabilitation in improving outcomes for patients with COVID-19.^38^ ^39^ The peak in the OE ratio occurred 1 month after the fifth wave, possibly attributed to the increased virulence of the delta variant. However, from October 2021 to February 2022, this trend shifted negatively, possibly owing to rationing measures or a decrease in patients who are severely ill as a result of vaccination and reduced virulence of the Omicron variant.^40–42^

The study found increased rehabilitation provisions per inpatient month across all three areas post-COVID-19. For cerebrovascular rehabilitation, two possibilities were considered: (1) decreased patient functionality because of limited rehabilitation and daily activities during the pandemic^43^ and (2) severe illness requiring intensive rehabilitation, possibly resulting from delays in emergency care and reduced availability of treatments such as mechanical thrombectomy.^44^ ^45^ However, some studies have reported no differences in stroke severity during the pandemic,^46–48^ highlighting the need for further research to fully understand these trends.

Our findings have important implications for policymakers. Monitoring the bed occupancy rate for infectious diseases can help estimate the impact of novel infectious diseases on rehabilitation services. Therefore, policymakers should use these rates to quantitatively measure their impact on rehabilitation services. Second, a quantitative assessment of risks and benefits is essential. This study suggests a trade-off between bed utilization for infectious diseases and rehabilitation services such as cerebrovascular and musculoskeletal services. During a pandemic, medical resources cannot be allocated to only one aspect of care. Policymakers can use quantitative indicators like quality-adjusted life-years to determine the appropriate balance. However, the calculation is complicated, requiring further research for rapid and timely assessment. Lastly, during the pandemic surges, bed shortages are inevitable; therefore, policymakers and healthcare providers should develop triage strategies to prioritize rehabilitation services.

Future research should develop strategies for maintaining access to rehabilitation services during health crises. Although the potential for online rehabilitation has been noted,^1^ ^49^ caution is warranted because of challenges such as limited Internet access for older adults and the need for tailored services.^2^ Long-term studies are required to assess the impact of such interventions on patient outcomes and healthcare system efficiency. Qualitative research is required to clarify the decision-making processes and healthcare delivery structures within individual medical institutions during bed shortages and patient surges. The healthcare system in crisis is a complex structure defined not only by the number of beds and healthcare workers but also by staff, resources, structures, and systems. ^50^ Decision-making during an emerging infectious disease outbreak often relies on limited information and differs from typical decision-making processes.^51^ Understanding these processes can help develop more effective strategies for maintaining essential services, including rehabilitation, during future health crises.

## Supporting information

RECORD Checklist

## Data Availability

The data set can be utilized for limited use, and its sharing with third parties is not allowed.

## List of abbreviations

COVID-19: coronavirus disease 2019
KDB: Kokuho Database System
OE: observed-to-expected
SARIMA: seasonal autoregressive integrated moving average

## Declarations

### Ethics approval and consent to participate

This study was approved by the Research Ethics Review Committee of the Graduate School of Health Innovation at Kanagawa University of Health and Welfare. All data were anonymized and handled in compliance with relevant data protection and privacy regulations. Patient informed consent was waived by the Research Ethics Review Committee of the Graduate School of Health Innovation at Kanagawa University of Health and Welfare as the data contains no individually identifying information.

### Consent for publication

Not applicable

### Availability of data and materials

The data set can be utilized for limited use, and its sharing with third parties is not allowed.

### Competing interests

The authors declare that they have no competing interests.

### Funding

Not applicable

### Author Contributions

Y.E. designed the study, the main conceptual ideas, and the proof outline. Y.E. and W.R. collected the data. Y.E. interpreted the results and worked on the manuscript. R.W. supervised the project. All authors discussed the results and commented on the manuscript.

## Notes

### Competing Interest Statement

The authors have declared no competing interest.

## References

1 Bettger JP, Thoumi A, Marquevich V, et al. COVID-19: maintaining essential rehabilitation services across the care continuum. BMJ Glob Heal. 2020;5:e002670. doi: 10.1136/bmjgh-2020-002670

2 Biase SD, Cook L, Skelton DA, et al. The COVID-19 rehabilitation pandemic. Age Ageing. 2020;49:696–700. doi: 10.1093/ageing/afaa118

3 Srinivas G, Maanasa R, Meenakshi M, et al. Ethical rationing of healthcare resources during COVID-19 outbreak: Review. Ethics Medicine Public Heal. 2021;16:100633. doi: 10.1016/j.jemep.2021.100633

4 Kapsner LA, Kampf MO, Seuchter SA, et al. Reduced Rate of Inpatient Hospital Admissions in 18 German University Hospitals During the COVID-19 Lockdown. Frontiers Public Heal. 2021;8:594117. doi: 10.3389/fpubh.2020.594117

5 Dufour E, Baheux C, Zureik M. Routine surgeries during the COVID-19 pandemic: A French nationwide cohort study. Ann Medicine Surg. 2022;77:103721. doi: 10.1016/j.amsu.2022.103721

6 Miyawaki A, Tomio J, Nakamura M, et al. Changes in Surgeries and Therapeutic Procedures During the COVID-19 Outbreak. Ann Surg. 2021;273:e132–4. doi: 10.1097/sla.0000000000004528

7 Mason LW, Malhotra K, Houchen-Wollof L, et al. The UK foot and ankle COVID-19 national (FAlCoN) audit – Regional variations in COVID-19 infection and national foot and ankle surgical activity. Foot Ankle Surg. 2022;28:205–16. doi: 10.1016/j.fas.2021.03.012

8 Wei Z, Xu Y, Feng B, et al. The impact of COVID-19 on hip and knee arthroplasty surgical volume in China. Int Orthop. 2023;1–8. doi: 10.1007/s00264-023-05944-1

9 Penfold CM, Blom AW, Redaniel MT, et al. The impact of restricted provision of publicly funded elective hip and knee joints replacement during the COVID-19 pandemic in England. PLOS ONE. 2023;18:e0294304. doi: 10.1371/journal.pone.0294304

10 Cieza A, Causey K, Kamenov K, et al. Global estimates of the need for rehabilitation based on the Global Burden of Disease study 2019: a systematic analysis for the Global Burden of Disease Study 2019. Lancet. 2021;396:2006–17. doi: 10.1016/s0140-6736(20)32340-0

11 WHO. Ageing and health. 2022. Available: https://www.who.int/news-room/fact-sheets/detail/ageing-and-health [Accessed 22 June 2024]

12 Taneda K, Kakinuma T, Nakanishi Y, et al. Community Health Care Vision Toward realizing the desired medical service system. J Natl Inst Public Heal. 2023;72:43–51. doi: 10.20683/jniph.72.1_43

13 Howard-Wilsher S, Irvine L, Fan H, et al. Systematic overview of economic evaluations of health-related rehabilitation. Disabil Heal J. 2016;9:11–25. doi: 10.1016/j.dhjo.2015.08.009

14 Briggs AM, Cross MJ, Hoy DG, et al. Musculoskeletal Health Conditions Represent a Global Threat to Healthy Aging: A Report for the 2015 World Health Organization World Report on Ageing and Health. Gerontol. 2016;56:S243–55. doi: 10.1093/geront/gnw002

15 Chen N, Fong DYT, Wong JYH. Secular Trends in Musculoskeletal Rehabilitation Needs in 191 Countries and Territories From 1990 to 2019. JAMA Netw Open. 2022;5:e2144198. doi: 10.1001/jamanetworkopen.2021.44198

16 Barman A, Sinha MK, Sahoo J, et al. Respiratory rehabilitation in patients recovering from severe acute respiratory syndrome: A systematic review and meta-analysis. Hear Lung. 2022;53:11–24. doi: 10.1016/j.hrtlng.2022.01.005

17 Jastrzebski D, Gumola A, Gawlik R, et al. Dyspnea and quality of life in patients with pulmonary fibrosis after six weeks of respiratory rehabilitation. J Physiol PharmacolJ: Off J Pol Physiol Soc. 2006;57 Suppl 4:139–48.

18 Economic Analysis of Respiratory Rehabilitation. J Cardiopulm Rehabilitation. 1998;18:238. doi: 10.1097/00008483-199805000-00013

19 Schindel D, Schneider A, Grittner U, et al. Quality of life after stroke rehabilitation discharge: a 12-month longitudinal study. Disabil Rehabilitation. 2021;43:2332–41. doi: 10.1080/09638288.2019.1699173

20 Maulden SA, Gassaway J, Horn SD, et al. Timing of Initiation of Rehabilitation After Stroke. Arch Phys Med Rehabilitation. 2005;86:34–40. doi: 10.1016/j.apmr.2005.08.119

21 Sahely A, Hew SKN, Chan YK, et al. Exploring the experiences of people who had a stroke and therapists who managed people with stroke during the COVID-19 pandemic: An exploratory qualitative study. PLOS ONE. 2023;18:e0282325. doi: 10.1371/journal.pone.0282325

22 Pandian JD, Kusuma Y, Kiemas LS, et al. Stroke Care During the COVID-19 Pandemic: Asian Stroke Advisory Panel Consensus Statement. J Stroke Med. 2021;4:7–14. doi: 10.1177/25166085211000915

23 Sousa DA de, Worp HB van der, Caso V, et al. Maintaining stroke care in Europe during the COVID-19 pandemic: Results from an international survey of stroke professionals and practice recommendations from the European Stroke Organisation. Eur Stroke J. 2020;5:230–6. doi: 10.1177/2396987320933746

24 National Health Service. COVID-19 Hospital Discharge Service Requirements. 2020. Available: https://www.gov.uk/government/publications/coronavirus-covid-19-hospital-discharge-service-requi <u>rements</u> [Accessed 8 September 2024]

25 Iuliano AD, Brunkard JM, Boehmer TK, et al. Trends in Disease Severity and Health Care Utilization During the Early Omicron Variant Period Compared with Previous SARS-CoV-2 High Transmission Periods — United States, December 2020–January 2022. Morb Mortal Wkly Rep. 2022;71:146–52. doi: 10.15585/mmwr.mm7104e4

26 United Nations. World Population Ageing 2019 Highlights. 2019. Available: https://www.un.org/development/desa/pd/sites/www.un.org.development.desa.pd/files/files/documents/2020/Jan/worldpopulationageing2019-highlights.pdf [Accessed 14 July 2024]

27 Kanazawa N, Inoue N, Tani T, et al. Implementation of Rehabilitation and Patient Outcomes During the Initial COVID-19 Pandemic. Prog Rehabilitation Med. 2022;7:20220031. doi: 10.2490/prm.20220031

28 Ministry of Health, Labour and Welfare. Overview of the 2022 Medical Facilities (Dynamic) Survey and Hospital Report (In Japanese). 2022. Available: https://www.mhlw.go.jp/toukei/saikin/hw/iryosd/22/ [Accessed 6 July 2024]

29 Narita N, Okumura K, Kinjo T, et al. Trends in prevalence of non-valvular atrial fibrillation and anticoagulation therapy in a Japanese region ― analysis using the national health insurance database ―. Circ J. 2020;84:706--713. doi: 10.1253/circj.cj-18-0989

30 Umebayashi R, Uchida HA, Matsuoka-Uchiyama N, et al. Prevalence of Chronic Kidney Disease and Variation of Its Risk Factors by the Regions in Okayama Prefecture. J Pers Med. 2022;12:97. doi: 10.3390/jpm12010097

31 Ministry of Health, Labour and Welfare. National Health Insurance Survey FY2022 Summary of Survey Results (In Japanese). 2022. Available: https://www.e-stat.go.jp/stat-search/files?page=1&layout=datalist&toukei=00450397&tstat=000001214140&cycle=8&tclass1=000001214141&result_page=1&tclass2val=0 [Accessed 4 July 2024]

32 Ministry of Health, Labour and Welfare. Survey on the state of medical care and the number of inpatient beds (In Japanese). Available: https://www.mhlw.go.jp/stf/seisakunitsuite/newpage_00023.html [Accessed 16 March 2022]

33 Kanagawa Prefectural Government. National Health Insurance Business Status [In Japanese]. Available: https://www.pref.kanagawa.jp/docs/n5p/cnt/f7093/p1128867.html [Accessed 24 October 2024]

34 Bolotov I. ARIMAAUTO: Stata module to find the best ARIMA model with the help of a Stata-adjusted Hyndman-Khandakar (2008) algorithm. 2022. Available: https://ideas.repec.org/c/boc/bocode/s459043.html [Accessed 6 July 2024]

35 Hung M, Mennell B, Christensen A, et al. Trends in COVID-19 Inpatient Cases and Hospital Capacities during the Emergence of the Omicron Variant in the United States. COVID. 2022;2:1207–13. doi: 10.3390/covid2090087

36 Chen DE, Tay EK, Tan PL, et al. The Impact of COVID-19 Pandemic on Rehabilitation in Singapore. *Ann Acad Med*, Singap. 2020;49:925–7. doi: 10.47102/annals-acadmedsg.2020297

37 Bestehorn K, Schwaab B, Schlitt A. Wie stark hat die COVID-19-Pandemie die kardiologische Rehabilitation im ersten Jahr der Pandemie beeinflusst? Ein Vergleich der Leistungszahlen aus 2019 mit 2020 in Deutschland. Z für Evidenz, Fortbild Qual im Gesundheitswesen. 2022;173:22–6. doi: 10.1016/j.zefq.2022.05.007

38 Nopp S, Moik F, Klok FA, et al. Outpatient Pulmonary Rehabilitation in Patients with Long COVID Improves Exercise Capacity, Functional Status, Dyspnea, Fatigue, and Quality of Life. Respiration. 2022;101:593–601. doi: 10.1159/000522118

39 AL-Mhanna SB, Mohamed M, Noor NM, et al. Effectiveness of Pulmonary Rehabilitation among COVID-19 Patients: A Systematic Review and Meta-Analysis. Healthcare. 2022;10:2130. doi: 10.3390/healthcare10112130

40 Zhao H, Lu L, Peng Z, et al. SARS-CoV-2 Omicron variant shows less efficient replication and fusion activity when compared with Delta variant in TMPRSS2-expressed cells. Emerg Microbes Infect. 2022;11:277–83. doi: 10.1080/22221751.2021.2023329

41 Esper FP, Adhikari TM, Tu ZJ, et al. Alpha to Omicron: Disease Severity and Clinical Outcomes of Major SARS-CoV-2 Variants. J Infect Dis. 2022;227:344–52. doi: 10.1093/infdis/jiac411

42 Kanagawa Prefectural Government. New coronavirus infection Kanagawa prefecture response record (In Japanese). 2023. Available: https://www.pref.kanagawa.jp/docs/ga4/covid19/archive/records.html [Accessed 9 September 2024]

43 Held JPO, Schwarz A, Pohl J, et al. Changes in stroke rehabilitation during the SARS-COV-2 shutdown in Switzerland. J Rehabilitation Med. 2022;54:1118. doi: 10.2340/jrm.v53.1118

44 Nogueira RG, Abdalkader M, Qureshi MM, et al. Global impact of COVID-19 on stroke care. Int J Stroke. 2021;16:573–84. doi: 10.1177/1747493021991652

45 Elgendy IY, Kumbhani DJ, Mahmoud A, et al. Mechanical Thrombectomy for Acute Ischemic Stroke A Meta-Analysis of Randomized Trials. J Am Coll Cardiol. 2015;66:2498–505. doi: 10.1016/j.jacc.2015.09.070

46 Jasne AS, Chojecka P, Maran I, et al. Stroke Code Presentations, Interventions, and Outcomes Before and During the COVID-19 Pandemic. Stroke. 2020;51:2664–73. doi: 10.1161/str.0000000000000347

47 Thau L, Siegal T, Heslin ME, et al. Decline in Rehab Transfers Among Rehab-Eligible Stroke Patients During the COVID-19 Pandemic. J Stroke Cerebrovasc Dis. 2021;30:105857. doi: 10.1016/j.jstrokecerebrovasdis.2021.105857

48 White TG, Martinez G, Wang J, et al. Impact of the COVID-19 Pandemic on Acute Ischemic Stroke Presentation, Treatment, and Outcomes. Stroke Res Treat. 2021;2021:8653396. doi: 10.1155/2021/8653396

49 Turolla A, Rossettini G, Viceconti A, et al. Musculoskeletal Physical Therapy During the COVID-19 Pandemic: Is Telerehabilitation the Answer? Phys Ther. 2020;100:pzaa093-. doi: 10.1093/ptj/pzaa093

50 Therrien M-C, Normandin J-M, Denis J-L. Bridging complexity theory and resilience to develop surge capacity in health systems. J Heal Organ Manag. 2017;31:96–109. doi: 10.1108/jhom-04-2016-0067

51 Rhodes R. Justice and Guidance for the COVID-19 Pandemic. Am J Bioeth. 2020;20:163–6. doi: 10.1080/15265161.2020.1777354

