## Supplementary Table 1 for "COVID-19 Bed Surge and Rationing of Rehabilitation Services in Japan: A Seasonal Autoregressive Integrated Moving Average (SARIMA) Analysis of Care Utilization with 10 years claims data"

Supplementary table 1: Parameters of SARIMA models

|  |  |  | SARIMA  model parameter | MAPE (%) | Dickey–Fuller test | White noise |
| --- | --- | --- | --- | --- | --- | --- |
| Service volume | | |  |  |  |  |
|  | Cerebral Vascular Rehabilitation | |  |  |  |  |
|  |  | Hospitalized | (1,0,0) (1,1,0,12) | 3.3 | * | *** |
|  |  | Outpatient | (0,0,1) (0,1,1,12) | 3.9 | ** | *** |
|  | Musculoskeletal Rehabilitation | |  |  |  |  |
|  |  | Hospitalized | (1,0,1) (0,1,1,12) | 3.5 | ** | *** |
|  |  | Outpatient | (1,0,1) (0,1,1,12) | 4.2 | ** | *** |
|  | Respiratory Rehabilitation | |  |  |  |  |
|  |  | Hospitalized | (0,0,1) (0,1,1,12) | 6.3 | ** | *** |
|  |  | Outpatient | (1,0,0) (1,1,0,12) | 11.4 | * | *** |
| Supplied patients' number | | |  |  |  |  |
|  | Cerebral Vascular Rehabilitation | |  |  |  |  |
|  |  | Hospitalized | (1,0,0) (1,1,0,12) | 2.7 | * | *** |
|  |  | Outpatient | (1,0,0) (1,1,0,12) | 2.3 | ** | *** |
|  | Musculoskeletal Rehabilitation | |  |  |  |  |
|  |  | Hospitalized | (1,0,1) (0,1,1,12) | 2.8 | ** | *** |
|  |  | Outpatient | (1,0,0) (1,1,0,12) | 2.0 | * | *** |
|  | Respiratory Rehabilitation | |  |  |  |  |
|  |  | Hospitalized | (1,0,0) (1,1,0,12) | 6.3 | ** | *** |
|  |  | Outpatient | (1,0,0) (1,1,0,12) | 11.4 | ** | *** |
|  |  |  |  |  |  | * p < 0.05 |
|  |  |  |  |  |  | ** p < 0.01 |
|  |  |  |  |  |  | *** p > 0.05 |
